# Race-Specific, U.S. State-Specific COVID-19 Vaccination Rates Adjusted for Age

**DOI:** 10.1101/2021.11.19.21266612

**Authors:** Elizabeth Wrigley-Field, Kaitlyn M. Berry, Govind Persad

## Abstract

We provide the first age-standardized race/ethnicity-specific, state-specific vaccination rates for the United States, encompassing all states reporting race/ethnicity-specific vaccinations. The data reflect vaccinations through mid-October 2021. We use indirect age standardization to compare racial/ethnic state vaccination rates to national age-specific vaccination patterns. Results show that white and Black state median vaccination rates are, respectively, 89% and 76% of what would be predicted based on age; Hispanic and Native rates are almost identical to what would be predicted; and Asian-American/Pacific Islander rates are 110% of what would be predicted. We also find that racial/ethnic group vaccination rates are associated with state politics, as proxied by 2020 Trump vote share: for each percentage point increase in 2020 Trump vote share, vaccination rates decline by 1.08 percent of what would be predicted based on age. This decline is sharpest for Native American populations, although Native vaccinations are reported for relatively few states.

---

In the United States, COVID-19 vaccination rates vary by race and by state (Ndugga et al. 2021). They also vary quite a bit by age, with 86% vaccination of U.S. residents aged 65-74, but only 53% vaccination of those aged 18-24, as of October 10, 2021 (see details in Appendix). This variation in vaccination rates by age can confound racial and state vaccination patterns because some population groups are older than others. In particular, the non-Hispanic white population is substantially older than all other populations: the median age in the U.S. is 43 for the non-Hispanic white population compared with 36 for the non-Hispanic Asian-American and Pacific Islander population, 33 for both non-Hispanic Black and Native populations, and just 30 for Hispanic populations. This suggests that aggregate population vaccination rates may be relatively higher for white populations compared to others simply because the white populations have more people at older ages.

COVID-19 mortality rates are frequently presented in age-adjusted form in order to facilitate comparisons across racial/ethnic and other populations (e.g., Ahmad et al. 2021), but vaccination rates are rarely presented this way--although one analysis examined age-adjusted vaccination rates for racial/ethnic groups in cities and towns in Massachusetts (Calef and Schuster 2021). Here, we present the first age-adjusted COVID-19 vaccination rates for each racial/ethnic group in each state in which data are available. These age-adjusted rates reveal racial and state vaccination patterns net of the shared pattern that older people are more likely than younger people to be vaccinated.

Our results capture the period shortly before vaccination was opened to ages 5-11. We combine Kaiser Family Foundation data on state-specific shares of vaccination going to each racial/ethnic group (released on October 18, 2021) with Centers for Disease Control and Prevention (CDC) data on state-level vaccination rates (released on October 21, 2021). Additionally, we incorporate subsidiary data collected by the CDC and National Center for Health Statistics (described in the Appendix).

We used a technique called “indirect age standardization” (Preston et al. 2001: 26-28) to create race-specific, state-specific vaccination levels that adjust for each group’s unique age distribution. For each racial group in each state, these indirectly standardized rates are the ratio of the actual vaccination rate to the vaccination rate that would be predicted solely based on that population’s age distribution (in conjunction with the national age pattern of COVID-19 vaccination). Thus, when this quantity exceeds 1, the group is more vaccinated than would be predicted based on age; when it is less than 1, it is less vaccinated than would be predicted. This method allows groups to be compared, revealing variation in their vaccination rates above and beyond that associated with age.

Results reveal two major patterns: (1) substantial racial variation in vaccination above and beyond the variation associated with age and (2) state-level variation, both overall and for all racial groups, that is associated with state politics.

Figure 1’s Panel A shows stark variation across racial groups in vaccination rates, adjusted for age. In most states, white and Black populations are vaccinated much less than would be expected based on their age distributions (with median adjusted vaccination rates across states of 89% and 76% of expectation, respectively); Hispanic and Native populations are vaccinated, on average, very similarly to what one would expect (median 100% and 99% of expectation, respectively); and Asian-American and Pacific Islander populations much more than would be expected (median 110% of expectation). The nearly flat smoothed curve of the Native distribution indicates vaccination rates that vary widely across states, without any clear peak range of values, though we caution that data on this population is reported by relatively few states (see Appendix).

**Figure 1.**
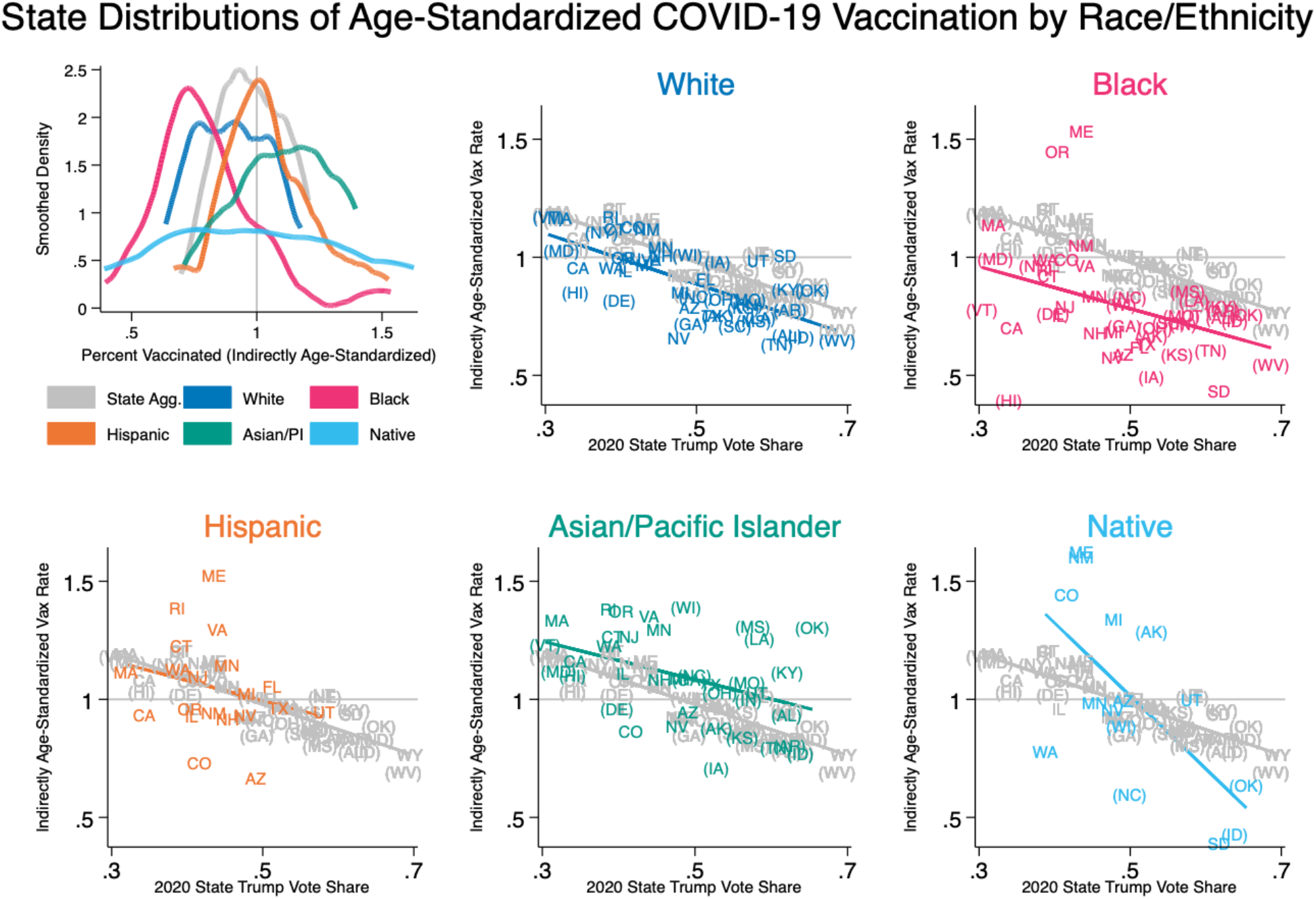
Age-standardized vaccination by race/ethnicity and state (as of October 2021) Indirectly age-standardized vaccination rates by state and race/ethnicity. When these rates exceed 1, the population displayed has vaccination rates higher than would be expected based on its age distribution; when the rates are less than 1, the population has vaccination rates lower than would be expected. Panel A shows the distribution across states of age-standardized vaccination rates for each racial/ethnic group, smoothed using the Epanechnikov kernel. Panels B-F show the state-specific, race/ethnicity-specific age-standardized vaccination rates as a function of states’ overall Trump vote share. Distributions in grey are state aggregate vaccination rates. States differ in their handling of Hispanic populations. Some exclude Hispanic individuals from other racial groups; others include Hispanic individuals in other racial groups. The latter states are labeled in Panels B-F with parentheses surrounding their state abbreviation for racial groups other than Hispanic, indicating that those racial groups include Hispanic individuals. Population denominators match states’ reported handling of Hispanic individuals in vaccination data. State-specific, race/ethnicity-specific vaccination rates are based on vaccine share data from the Kaiser Family Foundation (October 18, 2021 data release) and state-level vaccination rates from the CDC (October 21, 2021 data release). Age standardization is based on national age-specific vaccination rates from the CDC (October 15, 2021 data download, reflecting vaccinations through October 6, 2021) and state-specific, race/ethnicity-specific population estimates from the National Center for Health Statistics.

These vaccine distributions can also be weighted by population size--rather than treating each state equally--in order to better approximate national populations for each racial group rather than summarize states, although the distributions will not fully recover national populations since not all states report racial/ethnic data. When the distributions are weighted by population size, median age-adjusted vaccination rates are very similar to the medians of the state distributions for white and Black populations (91% and 75% of expectation, respectively). However, Hispanic and, especially, Native median vaccination rates are lower than when states are weighted equally (96% and 89% of expectation, respectively), indicating that Hispanic and Native people disproportionately live in states where they have low vaccination rates. Additionally, median Asian/Pacific Islander rates are even higher (117% of expectation), indicating that Asian/Pacific Islander people tend to live in states where they have particularly high vaccination rates.

Fig. 1 Panel A’s most striking pattern is that only a handful of states (Maine, Massachusetts, New Mexico, and Oregon, as well as Washington, D.C.) have Black populations whose vaccination rates are higher than one would predict based on national vaccination rates by age. This is particularly notable given the young age distribution of most states’ Black populations. Age adjustment in a young population will tend to make it look relatively more vaccinated by partially explaining low vaccination rates as simply the consequence of age. Yet even with this adjustment, Black vaccination rates are substantially lower than vaccination rates of all other racial groups.

The undervaccination of white populations, relative to expectation, is also an important result in Panel A. Age adjustment highlights the extent to which, net of age, white populations are less vaccinated than most others, which is partially disguised in aggregate population rates because the white populations are so much older than all others.

State-level COVID-19 vaccination rates are also associated with state politics, as shown in Fig. 1 Panels B-F, which use each state’s 2020 Trump vote share as a crude proxy for the state political context. Averaged across states, each additional percentage point of the state vote that went to Trump in 2020 is associated with the state’s total vaccination rate (adjusted for age) being lower by 1.08 percent of what it would be predicted to be based on the state’s age distribution. (All regression results are reported in the Appendix.) These results are consistent with other analyses showing that COVID-19 cases and deaths are strongly associated, at the county level, with partisan political leaning (Krieger et al. 2021).

This relationship between state politics and state vaccination levels is also reproduced for each separate racial group. (For Hispanic populations alone, the relationship between state Trump share and state vaccination rate is not statistically significant. Notably, these panels reveal that states with high Trump share do not report vaccination rates by Hispanic ethnicity, which also suggest that overall Hispanic vaccination distributions analyzed here may overreport vaccination. Some, but not all, states with high Trump share have very small Hispanic populations.) The available data do not have sufficient statistical power to distinguish the relationship of different racial groups’ vaccination to Trump share from one another. But as a simple description, we note that the white vaccination rate varies with Trump share to almost exactly the same degree as the state aggregate vaccination rates do, while Black, Hispanic, and Asian/Pacific Islander groups vary less. The Hispanic and Black groups’ age-adjusted vaccination have the smallest relationship to Trump vote, with a loss of only 0.89 (Black) or 0.85 (Hispanic) percentage points of expected vaccination associated with each additional point of Trump share. Strikingly, the Native rate of age-adjusted vaccination varies with Trump share far more dramatically than other racial groups’ rates do--with each additional Trump point associated with a 3.1 percentage point decline in Native vaccination relative to expectation. (However, results for Native populations should be interpreted cautiously, since those vaccinations are reported in the fewest states. See further data comments in the Appendix.)

We do not claim that Trump voting drives these vaccination rates. Rather, Trump vote share is used as a proxy for a broad range of state political characteristics that may be related to vaccination in many different ways, including via urbanicity, beliefs about COVID-19, and longtime patterns of funding and state support for public health infrastructure. Because lack of vaccination has multiple causes, the relatively lower vaccination rates in states with higher Trump support may also have different contributing factors for different racial groups living in those states. For example, it is possible that relatively low vaccination rates in the white populations disproportionately reflect self-consciously politicized vaccine rejection, while relatively low rates in other populations may be more likely to reflect characteristics of state health infrastructures.

These age-standardized rates reveal systematic and substantial variation in vaccination levels across states, racial groups, and the intersection of the two, net of differences in population age.

## Data Availability

All original data and all software code used to analyze them are available online at https://osf.io/h4p8t/

## Acknowledgements

The authors thank the Kaiser Family Foundation for answering questions about the data they have generously made available, and Michelle Niemann for feedback on an article draft. This research was supported by the Eunice Kennedy Shriver National Institute of Child Health and Human Development (NICHD) via the Minnesota Population Center (P2C HD041023) and via a fellowship to Kaitlyn M. Berry (F31 HD107980).

## Supplemental Methods Appendix to

This analysis integrates several distinct datasets, summarized and cited in Table S1. The most important dataset we draw on gives the portion of each state’s total vaccinations that were given to each racial/ethnic group (dataset 1 in Table S1), collected by the Kaiser Family Foundation (KFF). In order to transform these “vaccine shares” into age-standardized vaccination rates, there are two main steps. First, we transform vaccination shares into raw vaccination rates (i.e., per-capita vaccinations) for each racial/ethnic group in each state. Second, we standardize these raw rates for age using an “indirect age standardization” method. Here, we explain these two steps in detail.

Throughout, we use “vaccinated” to mean having received two doses of an mRNA vaccine or one dose of a Johnson and Johnson vaccine (i.e., “fully vaccinated” without consideration of booster shots). The time period we cover (with datasets reflecting cumulative vaccinations through early-to-mid October 2021) summarizes a situation in which people aged 12+ had been eligible for vaccination for long enough that appointments were widely available without delay in most places, but nobody younger than age 12 was yet eligible.

### Step 1: Constructing raw (crude) vaccination rates

No single dataset gives rates of vaccination by race/ethnicity for all states. In order to construct vaccination rates for each racial/ethnic group in each state, we took aggregate vaccination rates for each state (dataset 2) and applied the vaccine share for each racial/ethnic group (dataset 1) to those state aggregate rates. For example, if 50% of a state is vaccinated (dataset 2) and 10% of its vaccinations are of Hispanic people (dataset 1), then 5% of the state’s total population is vaccinated Hispanic people. We divided these by the portion of the state’s total population (dataset 3) that is Hispanic (datasets 4 and 5), which yielded the percentage of the state’s Hispanic population that is vaccinated. This percentage is what we refer to as a “raw rate” of vaccination for each racial/ethnic group in each state (also called a crude rate).

States vary widely in the apparent quality of their vaccination data (dataset 1), as summarized in Table S2. The data quality measures we report in Table S2 (e.g., percent of vaccinations with unknown race/ethnicity) are reported by KFF; the underlying vaccination data are collected by KFF from state departments of health. We drop Pennsylvania from all analyses because Pennsylvania’s reported race/ethnicity-specific vaccination data (dataset 1) exclude Philadelphia, while Pennsylvania’s aggregate vaccination rates (dataset 2) and populations (datasets 3-5) include Philadelphia.

### Step 2: Age-standardizing vaccination rates

There are two ways to age-standardize rates. “Direct age standardization” involves reweighting age-specific rates for each population. However, age-specific vaccination rates are not available for race/ethnicity-specific, state-specific populations. Therefore, we use “indirect age standardization.” This method translates each racial/ethnic state population’s vaccination rate into a scale reflecting the degree to which the population is more vaccinated or less vaccinated than would be expected, based on the population’s age distribution.

In order to estimate these indirectly age-standardized rates, we begin with a set of age-specific vaccination rates for the United States as a whole (dataset 6). This age pattern is shown in Figure S1 across the Centers for Disease Control and Prevention (CDC)’s age categories (ages <12, 12-15, 16-17, 18-24, 25-39, 40-49, 50-64, 65-74, and 75+), which are used throughout the analysis. For comparison, Figure S2 shows the distribution across all US states of age for each racial/ethnic group. Note that the CDC vaccination rates for ages 65-74 seem unrealistically high, which may reflect inaccurate population denominators and/or booster shots (perhaps among those not yet eligible) being misreported as new vaccinations. We use a measure of “fully-vaccinated” rather than “any vaccine” (measuring having received at least one dose) in order to limit the latter possibility. Note also that, although nobody younger than age 12 was formally eligible, the CDC data (dataset 6) do show some vaccinations among younger children (0.3% vaccination among children under age 12). Since all racial/ethnic state populations will be normalized to this standard age schedule of vaccination, the comparative analysis will be unbiased as long as any errors in the CDC age schedule of vaccination are proportional across ages but will be biased if the CDC data contain greater error for some ages compared with others.

Using this national age pattern of vaccination and the age distribution of each racial/ethnic group population in each state (datasets 4 and 5), we estimate the “expected” vaccination rate of each racial/ethnic group in each state based on its age distribution. This reflects the aggregate vaccination rate that each state would have if it had the national aggregate vaccination rates for each of its age groups.

Finally, we take the ratio of the actual vaccination rate to this expected vaccination rate for each racial/ethnic population in each state. This quantity is sometimes called a Comparative Mortality Ratio (CMR). When the CMR exceeds 1, it means the population is more vaccinated than would be expected; for example, a CMR of 1.2 means a population whose raw vaccination rate is 120% of what would be expected based on its age distribution. When the CMR is less than 1, the population is less vaccinated than would be expected. This CMR is the outcome for each analysis shown in Figure 1.

### Racial/ethnic groups

The racial categories we use are white, Black, Asian-American/Asian/Pacific Islander/Native Hawaiian, Native, and Hispanic. The KFF vaccination dataset (dataset 1) distinguishes Asian from native Hawaiian, but because the National Center for Health Statistics (NCHS) population data (datasets 3-5) use a collapsed Asian/PI category that includes native Hawaiians, we collapse these categories in the vaccination rates as well.

Not all states report each racial/ethnic group. These states are listed, along with other limitations of dataset 1, in Table S2. We use all reported racial/ethnic categories in each state, except for “other race.” We omit this category (except for Hispanic individuals in this category, who are treated as Hispanic) out of concern that this group may have a different meaning in state vaccination records than in NCHS population denominators. In addition, the racial shares that we use are shares among vaccinations with known race/ethnicity. In some states, the percent of vaccinations with unknown race/ethnicity is quite high--up to 21% (in Washington, DC).

States differ in their treatment of Hispanic populations. Some states treat “Hispanic” as a racial group. In these states, “White” means non-Hispanic White, and so forth. We refer to these as states with “mutually exclusive” categories. Other states treat Hispanic as a “cross-cutting group.” In those states, “White” includes White Hispanic, and the total vaccination shares add up to more than 100% because the Hispanic population shares are double-counted. We relied on KFF data notes to distinguish how each state handled Hispanic populations (checked against whether vaccination shares total to 100%, with rounding error, or more than 100%).

We constructed population denominators that match each way of counting the Hispanic population: mutually exclusive (dataset 4) or cross-cutting (dataset 5). We used the appropriate denominator for each state so that vaccinations and populations were constructed in the same way. The handling of Hispanic populations in constructing population denominators can matter substantively.

In Figure 1’s Panels B-D, state abbreviations for non-Hispanic populations are denoted with parentheses when the state uses cross-cutting Hispanic populations and without parentheses when the state uses mutually exclusive Hispanic populations. When states report racial groups with cross-cutting Hispanic populations (the groups denoted with parentheses), race-specific divergences from what would be expected based on age may be understated, compared to the divergences of mutually exclusive racial groups, because the Hispanic populations (where reported) have vaccinations close to what would be expected based on their age distributions.

### Analyses

The density distributions shown in Figure 1 Panel A are estimated with Epanechnikov kernels with bandwidth chosen to minimize mean integrated squared error under assumption of a Gaussian distribution and kernel, estimated using Stata.

In our analysis of vaccination rates in relation to 2020 Trump vote share (dataset 7), we treat Trump share as a broad proxy for a wide array of state politics, which this analysis is unable to distinguish. This analysis omits Washington, D.C., which is included elsewhere as a “state” (in Figure 1 Panel A), because its Trump vote share (5%) is far outside the range of all other states (30%-69%). Regression results are reported in Table S3 and Table S4.

### Repository

Data and Stata code can be accessed at https://osf.io/h4p8t/

## APPENDIX FIGURES

**Figure S1.**
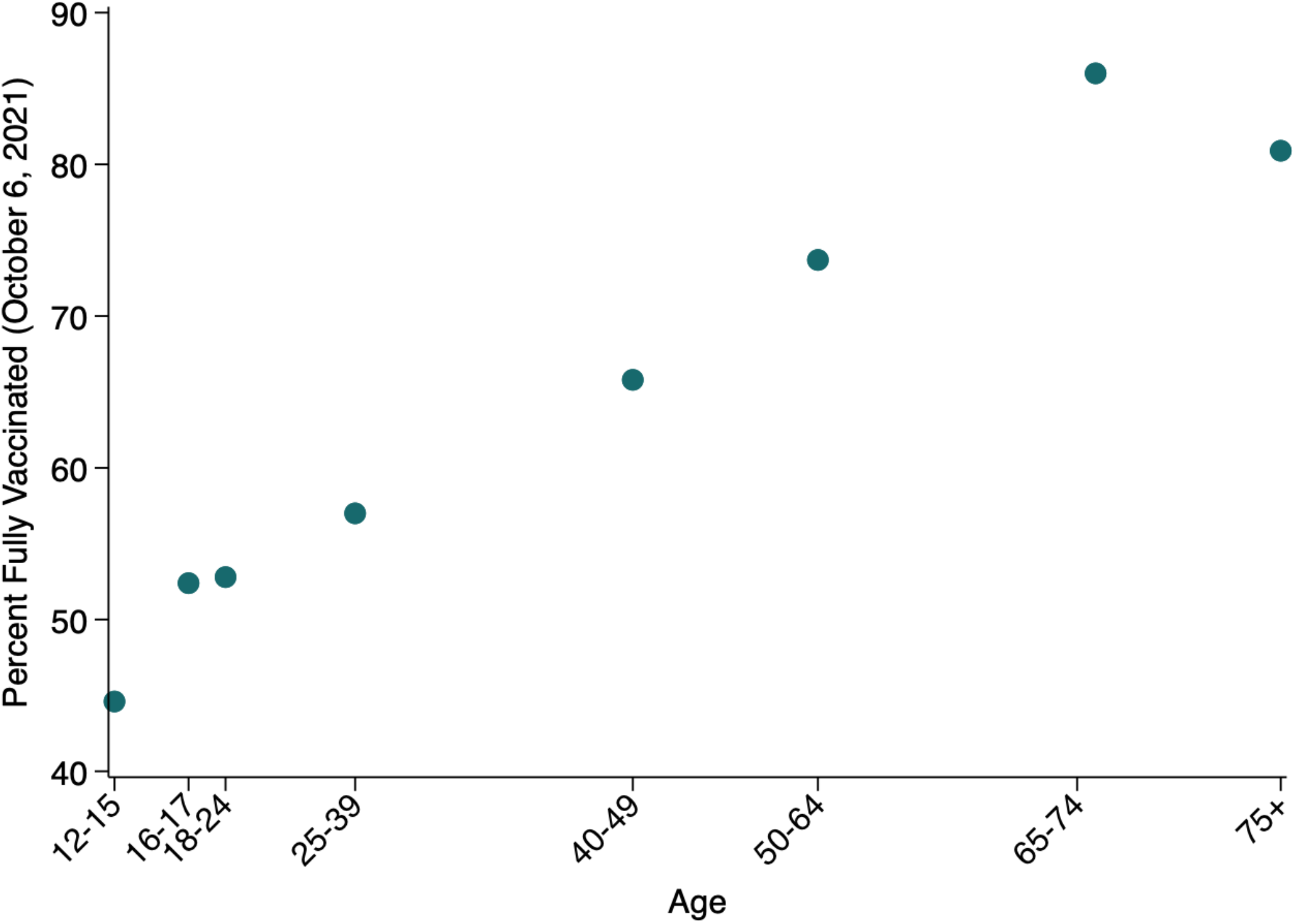
Vaccination status by age. Data: CDC (Table S1’s “dataset 6”).

**Figure S2.**
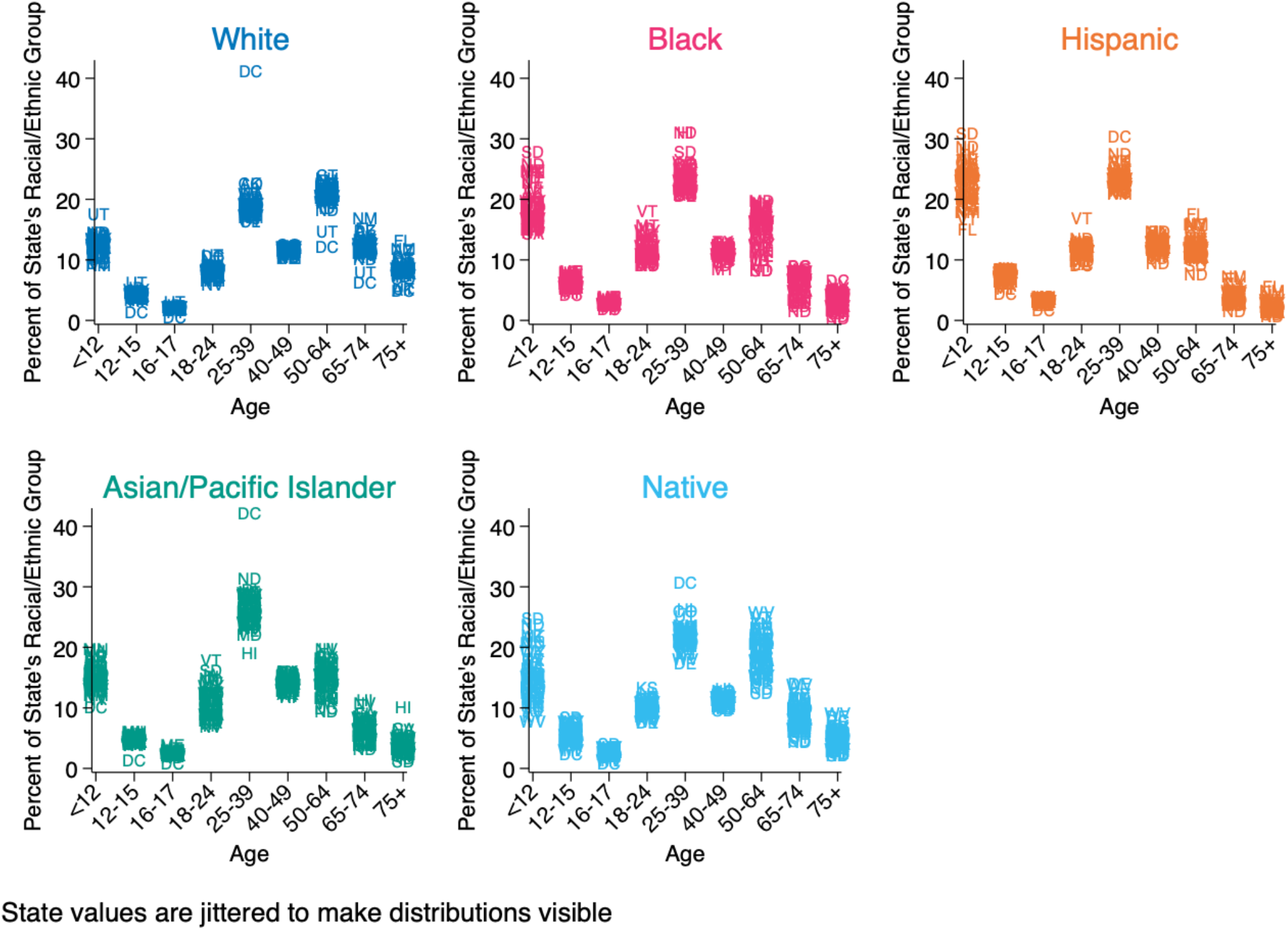
Age distributions by race and state. Data: NCHS (Table S1’s “dataset 4”). All racial groups besides Hispanic are limited to non-Hispanic individuals.

## APPENDIX TABLES

**Table S1.** Datasets used in analysis

| # | Description | Source | Date Downloaded | Citation |
| --- | --- | --- | --- | --- |
| 1 | Vaccine share for each racial/ethnic group | KFF | 10/18/2021 | https://github.com/KFFData/COVID-19-Data/tree/kff_master/Race%20Ethnicity%20COVID-19%20Data/Vaccines |
| 2 | Aggregate vaccination rates for each state | CDC | 10/21/2021 | https://covid.cdc.gov/covid-data-tracker/#vaccinations_vacc-total-admin-rate-total |
| 3 | State populations | NCHS | 11/3/2021 | https://wonder.cdc.gov/controller/saved/D178/D245F115 |
| 4 | Race-specific, age-specific state populations where Hispanic is a mutually exclusive category | NCHS | 10/22/2021 | https://wonder.cdc.gov/controller/saved/D178/D241F020 |
| 5 | Race-specific, age-specific state populations where Hispanic crosscuts racial categories | NCHS | 10/22/2021 | https://wonder.cdc.gov/controller/saved/D178/D241F020 |
| 6 | Age-specific national vaccination rates | CDC | 10/15/2021 | https://data.cdc.gov/Vaccinations/COVID-19-Vaccination-and-Case-Trends-by-Age-Group-/gxj9-t96f |
| 7 | State 2020 Trump vote share | MEDSL | 9/28/2021 | https://dataverse.harvard.edu/dataset.xhtml?persistentId=doi:10.7910/DVN/42MVDX |

KFF = Kaiser Family Foundation
CDC = Centers for Disease Control and Prevention
NCHS = National Center for Health Statistics
MEDSL = MIT Election Data and Science Lab
Dataset #4 and Dataset #5 are derived from the same underlying NCHS dataset, as cited above, by the research team.

**Table S2.** Data limitations in state vaccination data (Dataset 1)

| **State** | **Any race-specific vaccine data** | **Racial groups with at least 1% total vaccination share** | **Hispanic crosscuts racial categories** | **Percent of vaccinations attributed to “other race”** | **Percent of vaccinations with unknown race** |
| --- | --- | --- | --- | --- | --- |
| Alabama | Yes | white, Asian, Black, Hispanic | Yes | 11% | 11% |
| Alaska | Yes | white, Asian, Hawaiian/Pacific Islander, Black, Hispanic, Native | Yes | 12% | 8% |
| Arizona | Yes | white, Asian, Black, Hispanic, Native | No | 18% | 11% |
| Arkansas | Yes | white, Asian, Black, Hispanic | Yes | 8% | 7% |
| California | Yes | white, Asian, Hawaiian/Pacific Islander, Black, Hispanic | No | 11% | 5% |
| Colorado | Yes | white, Asian, Black, Hispanic, Native | No | 4% | 10% |
| Connecticut | Yes | white, Asian, Black, Hispanic | No | 7% | 5% |
| Delaware | Yes | white, Asian, Black, Hispanic | Yes | 21% | 3% |
| District of Columbia | Yes | white, Asian, Black, Hispanic | Yes | . | 21% |
| Florida | Yes | white, Black, Hispanic | No | 13% | 13% |
| Georgia | Yes | white, Asian, Black, Hispanic | Yes | 14% | 5% |
| Hawaii | Yes | white, Asian, Hawaiian/Pacific Islander, Black | Yes | 3% | 19% |
| Idaho | Yes | white, Asian, Black, Hispanic, Native | Yes | 15% | 18% |
| Illinois | Yes | white, Asian, Black, Hispanic | No | 4% | 4% |
| Indiana | Yes | white, Asian, Black, Hispanic | Yes | 11% | 4% |
| Iowa | Yes | white, Asian, Black, Hispanic | Yes | 2% | 10% |
| Kansas | Yes | white, Asian, Black, Hispanic | Yes | 19% | 9% |
| Kentucky | Yes | white, Asian, Black | Yes | 10% | 6% |
| Louisiana | Yes | white, Asian, Black, Hispanic | Yes | 7% | 14% |
| Maine | Yes | white, Asian, Black, Hispanic, Native | No | 16% | 6% |
| Maryland | Yes | white, Asian, Black, Hispanic, Native | Yes | 10% | 5% |
| Massachusetts | Yes | white, Asian, Black, Hispanic | No | 3% | NR |
| Michigan | Yes | white, Asian, Black, Hispanic, Native | No | 9% | 16% |
| Minnesota | Yes | white, Asian, Black, Hispanic, Native | No | 1% | 7% |
| Mississippi | Yes | white, Asian, Black, Hispanic | Yes | 3% | NR |
| Missouri | Yes | white, Asian, Black, Hispanic | Yes | . | NR |
| Montana | No |  | No | . | NR |
| Nebraska | No |  | Yes | . | NR |
| Nevada | Yes | white, Asian, Black, Hispanic, Native | No | 19% | NR |
| New Hampshire | Yes | white, Asian, Black, Hispanic | No | 4% | 8% |
| New Jersey | Yes | white, Asian, Black, Hispanic | No | 13% | 8% |
| New Mexico | Yes | white, Asian, Black, Hispanic, Native | No | 4% | 11% |
| New York | Yes | white, Asian, Black, Hispanic | Yes | 2% | NR |
| North Carolina | Yes | white, Asian, Black, Hispanic, Native | Yes | 8% | 6% |
| North Dakota | No |  | Yes | . | NR |
| Ohio | Yes | white, Asian, Black, Hispanic | Yes | 9% | 4% |
| Oklahoma | Yes | white, Asian, Black, Hispanic, Native | Yes | 5% | 19% |
| Oregon | Yes | white, Asian, Hawaiian/Pacific Islander, Black, Hispanic, Native | No | 4% | 7% |
| Pennsylvania | Yes | white, Asian, Black, Hispanic | Yes | 14% | 10% |
| Rhode Island | Yes | white, Asian, Black, Hispanic | No | . | NR |
| South Carolina | Yes | white, Black, Hispanic | Yes | 13% | 7% |
| South Dakota | Yes | white, Black, Native | No | 3% | 9% |
| Tennessee | Yes | white, Asian, Black, Hispanic | Yes | 24% | 4% |
| Texas | Yes | white, Asian, Black, Hispanic | No | 15% | 6% |
| Utah | Yes | white, Asian, Hawaiian/Pacific Islander, Black, Hispanic, Native | No | 3% | 13% |
| Vermont | Yes | white, Asian, Black, Hispanic | Yes | 2% | 3% |
| Virginia | Yes | white, Asian, Black, Hispanic | No | 6% | 8% |
| Washington | Yes | white, Asian, Hawaiian/Pacific Islander, Black, Hispanic, Native | No | 11% | 7% |
| West Virginia | Yes | white, Black | Yes | 8% | 6% |
| Wisconsin | Yes | white, Asian, Black, Hispanic, Native | Yes | 9% | 5% |
| Wyoming | No |  | No | . | NR |

NR=Not reported. Source for all data: Kaiser Family Foundation (Table S1's "dataset 1").

**Table S3.**
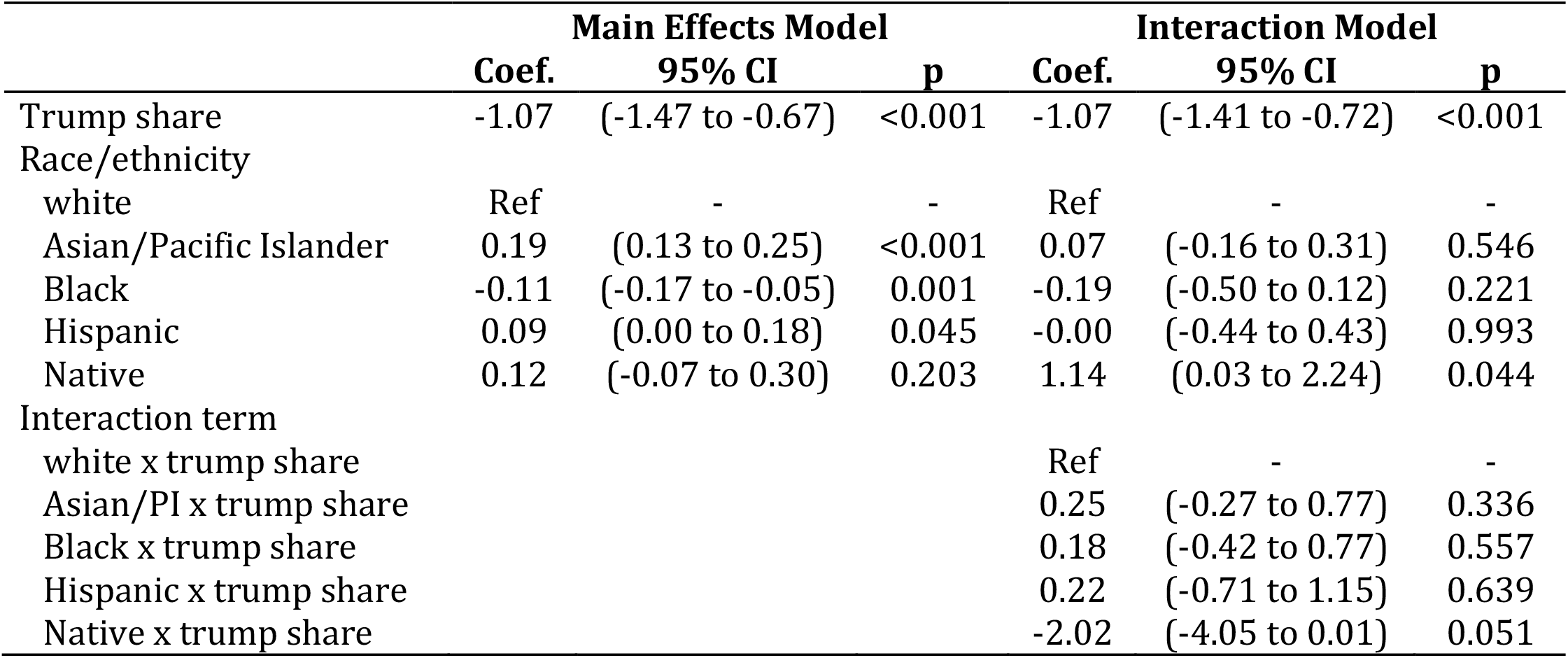
State age-standardized vaccination rates (aggregated over race) as a function of Trump vote share

**Table S4.** Race/ethnicity-specific state age-standardized vaccination rates as a function of Trump vote share

| Model | Included race/ethnicity | Coefficient for Trump Share |  |  |
| --- | --- | --- | --- | --- |
|  |  | Coef. | 95% CI | p-value |
| 1 | white | -1.07 | (-1.41 to -0.72) | <0.001 |
| 2 | Asian/Pacific Islander | -0.82 | (-1.40 to -0.23) | 0.008 |
| 3 | Black | -0.89 | (-1.50 to -0.28) | 0.005 |
| 4 | Hispanic | -0.85 | (-2.43 to 0.74) | 0.276 |
| 5 | Native | -3.09 | (-5.30 to -0.87) | 0.010 |

Each model shows state age-standardized vaccination rates as a function of Trump share for a single racial/ethnic group. For example, Model 1 only includes the age-standardized vaccination rates for the white population in each state.

